# Performance of ChatGPT and GPT-4 on Neurosurgery Written Board Examinations

**DOI:** 10.1101/2023.03.25.23287743

**Authors:** Rohaid Ali, Oliver Y. Tang, Ian D. Connolly, Patricia L. Zadnik Sullivan, John H. Shin, Jared S. Fridley, Wael F. Asaad, Deus Cielo, Adetokunbo A. Oyelese, Curtis E. Doberstein, Ziya L. Gokaslan, Albert E. Telfeian

**Affiliations:** Department of Neurosurgery, The Warren Alpert Medical School of Brown University, Providence, RI, USA; Department of Neurosurgery, University of Pittsburgh, Pittsburgh, PA, USA; Department of Neurosurgery, Massachusetts General Hospital, Boston, MA, USA

**Keywords:** Neurosurgery, medical education, surgical education, residency education, artificial intelligence, large language models, ChatGPT, GPT-4

## Abstract

**Background:** Interest surrounding generative large language models (LLMs) has rapidly grown. While ChatGPT (GPT-3.5), a general LLM, has shown near-passing performance on medical student board examinations, the performance of ChatGPT or its successor GPT-4 on specialized exams and the factors affecting accuracy remain unclear.

**Objective:** To assess the performance of ChatGPT and GPT-4 on a 500-question mock neurosurgical written boards examination.

**Methods:** The Self-Assessment Neurosurgery Exams (SANS) American Board of Neurological Surgery (ABNS) Self-Assessment Exam 1 was used to evaluate ChatGPT and GPT-4. Questions were in single best answer, multiple-choice format. Chi-squared, Fisher’s exact, and univariable logistic regression tests were employed to assess performance differences in relation to question characteristics.

**Results:** ChatGPT (GPT-3.5) and GPT-4 achieved scores of 73.4% (95% confidence interval [CI]: 69.3-77.2%) and 83.4% (95% CI: 79.8-86.5%), respectively, relative to the user average of 73.7% (95% CI: 69.6-77.5%). Question bank users and both LLMs exceeded last year’s passing threshold of 69%. While scores between ChatGPT and question bank users were equivalent (*P*=0.963), GPT-4 outperformed both (both *P*<0.001). GPT-4 answered every question answered correctly by ChatGPT and 37.6% (50/133) of remaining incorrect questions correctly. Among twelve question categories, GPT-4 significantly outperformed users in each but performed comparably to ChatGPT in three (Functional, Other General, and Spine) and outperformed both users and ChatGPT for Tumor questions. Increased word count (odds ratio [OR]=0.89 of answering a question correctly per +10 words) and higher-order problem-solving (OR=0.40, *P*=0.009) were associated with lower accuracy for ChatGPT, but not for GPT-4 (both *P*>0.005). Multimodal input was not available at the time of this study so, on questions with image content, ChatGPT and GPT-4 answered 49.5% and 56.8% of questions correctly based upon contextual context clues alone.

**Conclusion:** LLMs achieved passing scores on a mock 500-question neurosurgical written board examination, with GPT-4 significantly outperforming ChatGPT.

## Introduction

Artificial intelligence (AI) systems promise many potential applications in medicine, such as differential diagnosis generation and selection, clinical decision support, and analysis of imaging-, physiologic-, and genomic-based data.^1,2^ Within this discipline, attention has grown around ChatGPT (OpenAI; San Francisco, CA), a general Large Language Model developed by OpenAI and initially launched for public use in November 2022. ChatGPT, also known as GPT-3.5, was trained on a large corpus of text data through a combination of supervised and unsupervised learning techniques, followed by fine-tuning via reinforcement learning with human feedback. Notably, ChatGPT functions as an isolated language model that is incapable of searching the Internet, in contrast to other chatbots that can access external data. While OpenAI’s internal version of ChatGPT can query the Internet and these functions will likely be implemented in future public releases, the publicly available ChatGPT model does not presently have these capabilities. On March 14, 2023, OpenAI released an updated LLM entitled GPT-4, which was trained using a similar methodology as its predecessor.^3^ Moreover, GPT-4 notably introduced multimodal capabilities, such as the ability to input images, although these functions had yet to be released for public use at the time of this study.

Given this consideration and ongoing attention on the ability of AI models to supplement clinician knowledge and decision-making, the performance of systems like ChatGPT and GPT-4 on clinical board examinations has emerged as an area of intense interest. Kung *et al*. recently determined that ChatGPT approached a passing score on the United States Medical Licensing Examination (USMLE) Step 1 examination,^4^ a test traditionally administered after two years of preclinical education for which the average medical student studies approximately 400 hours.^5^ The model also performed similarly on the USMLE Step 2 and Step 3 examinations, scoring at >50% accuracy across all three. GPT-4 has additionally demonstrated performance improvements in other standardized exams, relative to ChatGPT (GPT-3.5), achieving a passing score in over 25 examinations across multiple disciplines. Most notably, while ChatGPT scored at the 10^th^ percentile for a mock bar examination, GPT-4 scored in the 90^th^ percentile.^3^ Moreover, GPT-4 has also demonstrated an over 20% improvement in all three USMLE Step examinations.^6^

While the USMLE examinations represent a holistic assessment of medical knowledge, the performance of LLMs has yet to be evaluated for more specialized medical board examinations, including in the setting of neurosurgery. Moreover, it is poorly understood if the performance of ChatGPT and GPT-4 is modulated by question characteristics such as length, subspecialty area, and incorporation of higher-order problem solving skills, in contrast to first-order recall. Consequently, the goal of this present study was to elucidate the performance of ChatGPT and GPT-4 on a mock neurosurgical written board examination.

## Methods

Performance of ChatGPT and GPT-4 was evaluated using the 500-question Self-Assessment Neurosurgery Exams (SANS) American Board of Neurological Surgery (ABNS) Self-Assessment Exam 1. Each question was entered individually in a single best answer multiple-choice format, with the original question and answer choices reproduced verbatim (**Figure 1A-E**). Because ChatGPT accepts exclusively text input and the multimodal capabilities of GPT-4 were not yet publicly available, no image data was provided as input to either model. Nonetheless, questions with images were used in this study by providing as input only the text portion.

**Figure 1:**
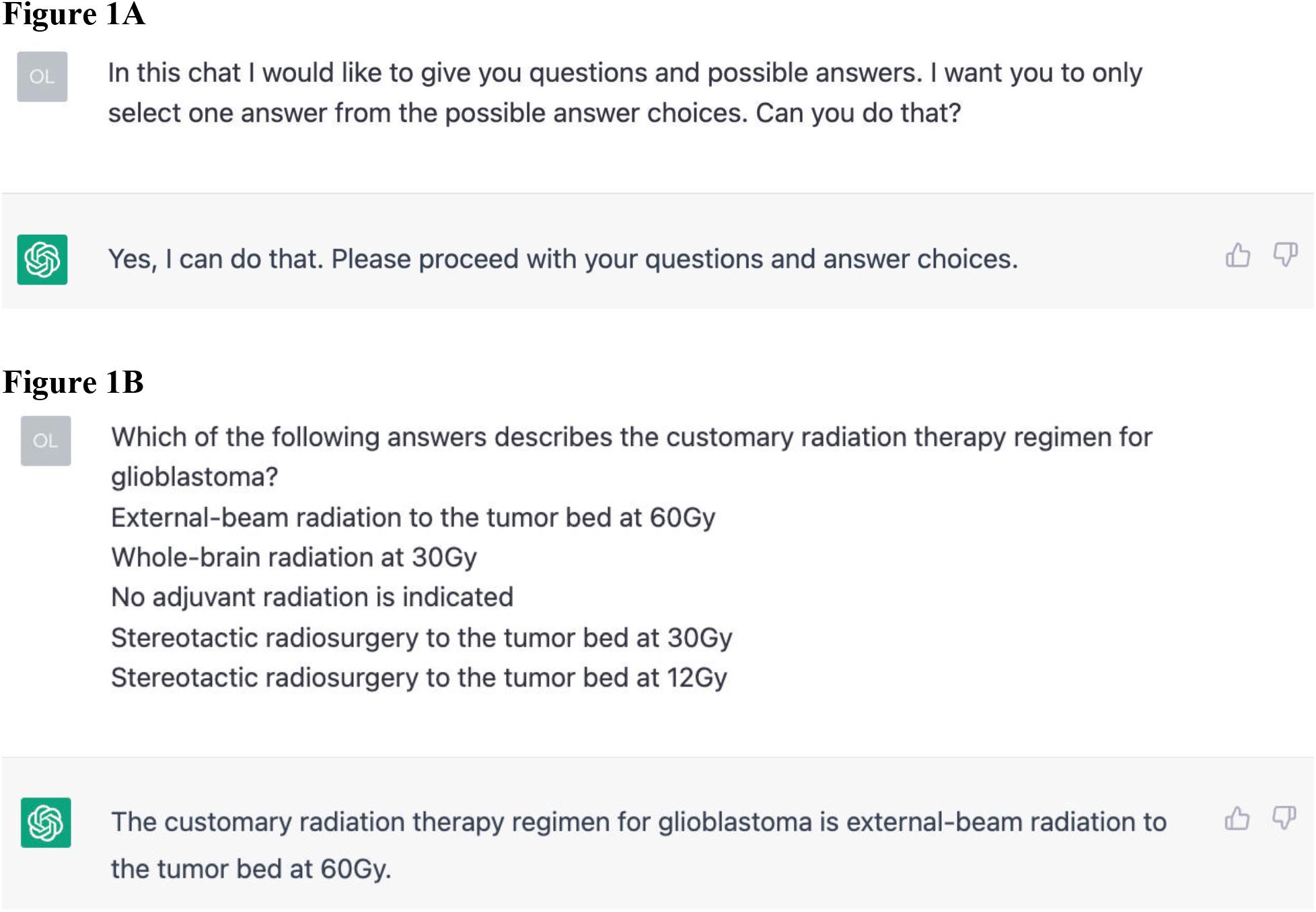
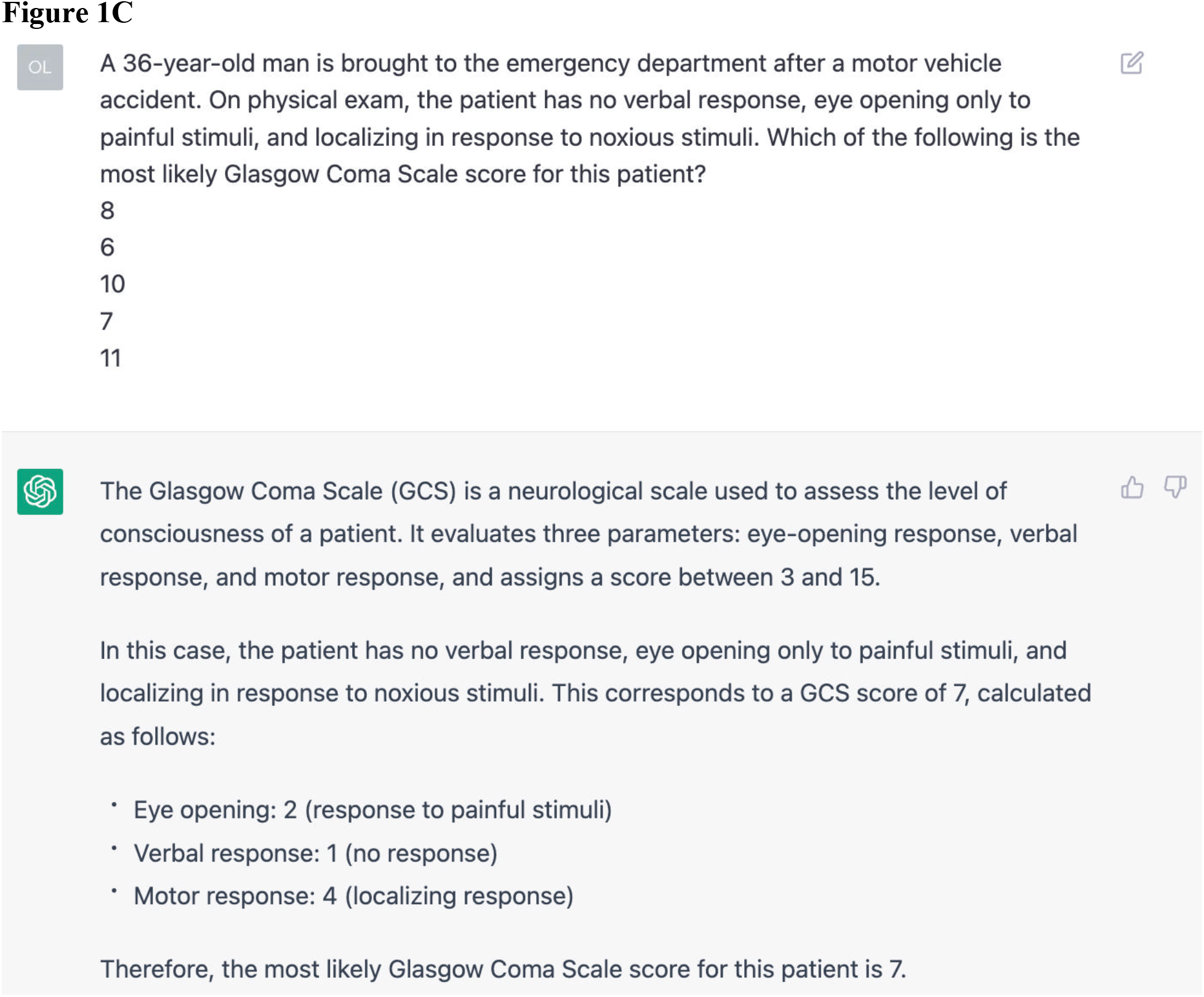
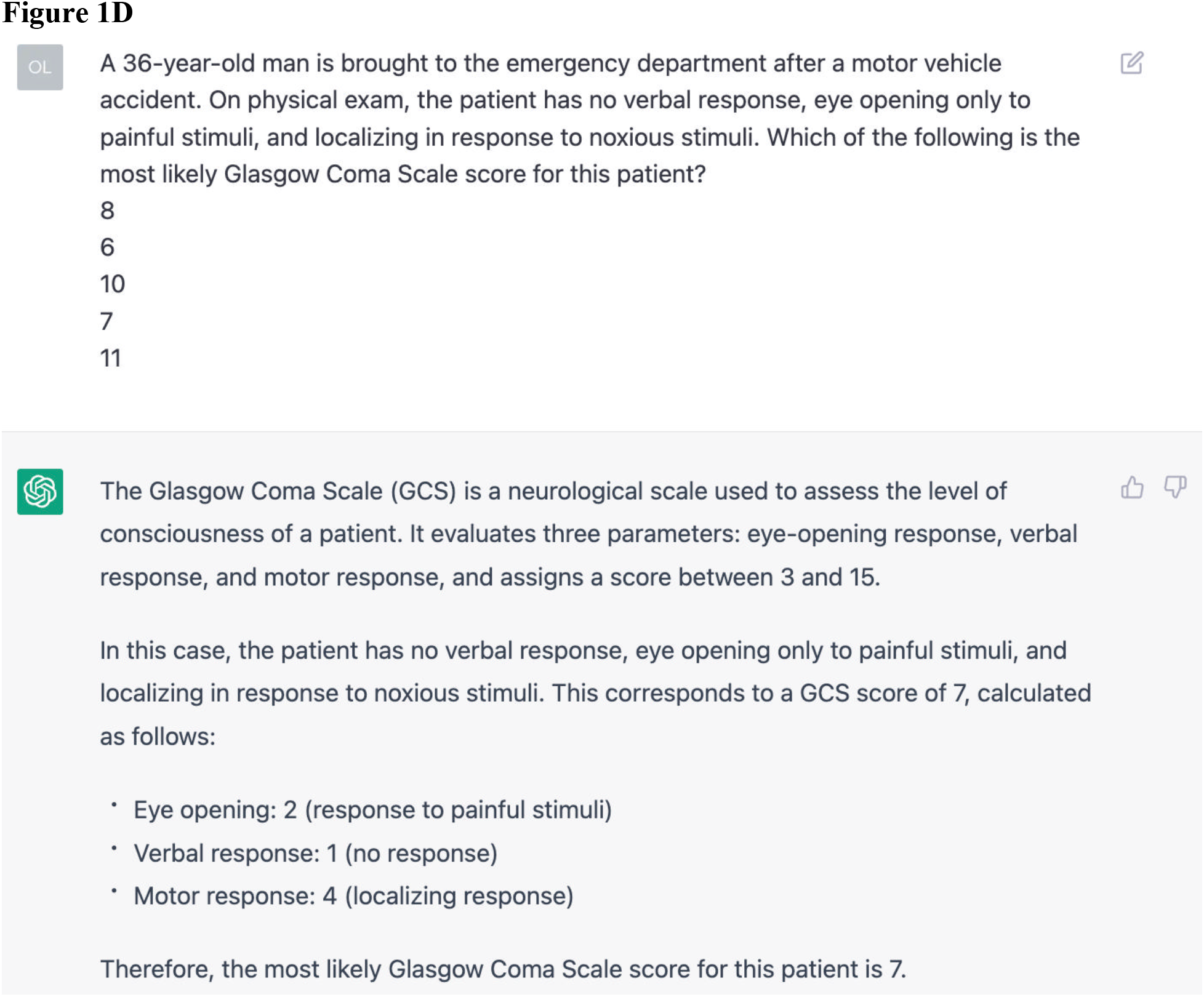
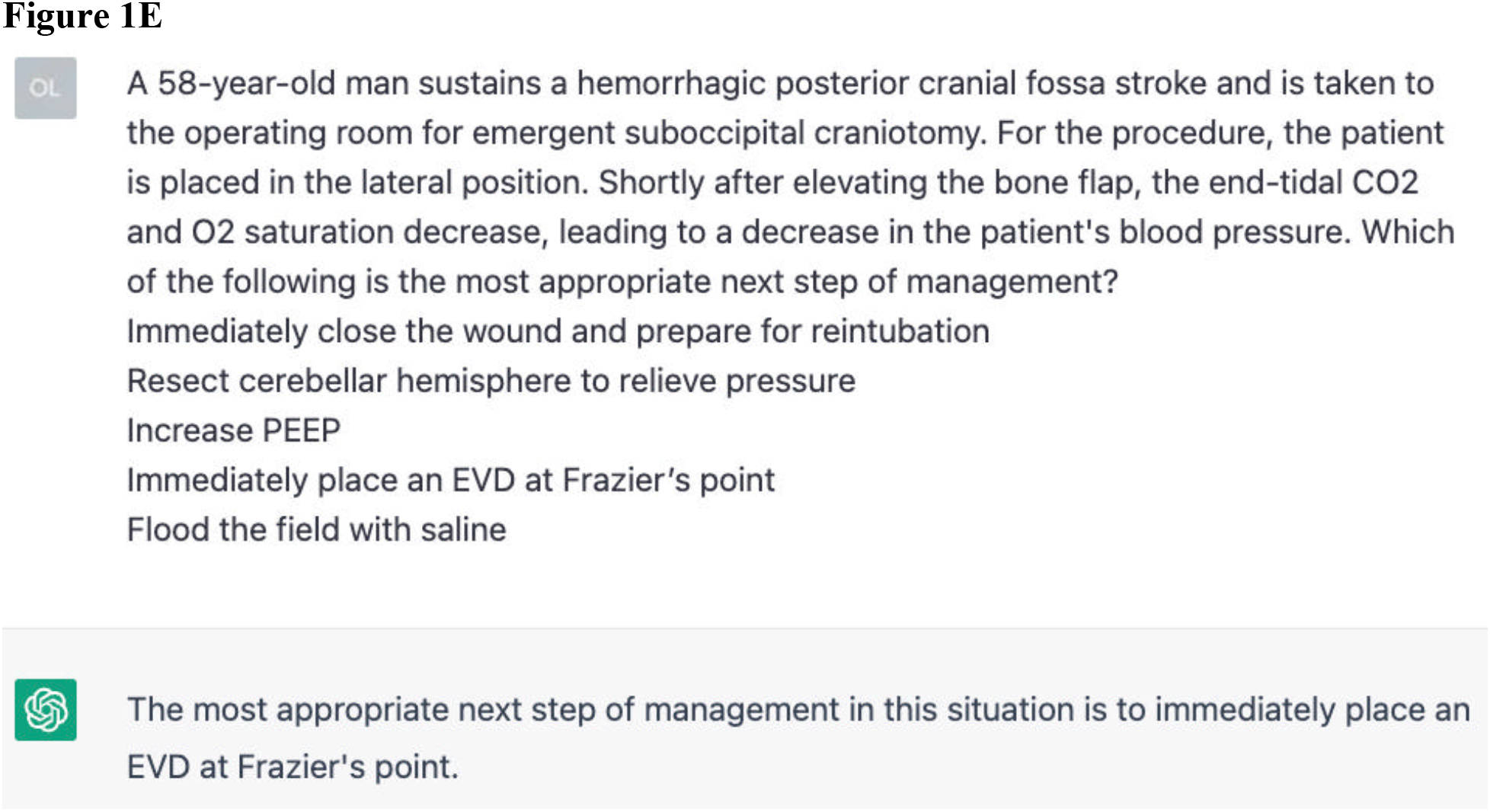
Representative Correct and Incorrect Questions Answered by ChatGPT (GPT-3.5) Screenshots of four questions answered by ChatGPT, illustrating the prompt used to input questions and sample responses. All images are attributed to ChatGPT (OpenAI; San Francisco, CA). **A:** Prompt for chat, used to request answers to be returned in a multiple single choice format. This prompt was used prior to inputting questions where ChatGPT did not return a single best answer choice. **B:** Correct answer for first-order question on adjuvant radiotherapy for glioblastoma. **C:** Incorrect answer for first-order question on calculating a Glasgow Coma Scale (GCS) score. To illustrate the underlying rationale (ChatGPT mistakenly and repeatedly referred to localizing pain as 4 points, rather than 5), a screenshot of ChatGPT’s response without prompting for a single multiple-choice answer is provided. **D:** Correct answer for higher-order question on the best surgical approach for a single-level lumbar disc herniation. To illustrate the underlying rationale, a screenshot of ChatGPT’s response without prompting for a single multiple-choice answer is provided. **E:** Incorrect answer for higher-order question on managing an intraoperative venous air embolism. Upon requesting a rationale for the answer (not shown), ChatGPT mistakenly diagnosed the etiology of the symptoms as elevated intracranial pressure.

Questions were already classified into one of twelve possible categories by test writers. User (neurosurgery trainee) performance by question category was collected from the test portal, but performance on individual-level questions was not reported. With manual evaluation, questions were also independently classified by two authors (RA and OYT) as incorporating first-order or higher-order problem-solving (**Table 1**). First-order questions were defined as those involving simple fact recall, such as identifying the mechanism of action of a medication or, most commonly, selecting the most likely diagnosis for a clinical vignette. Higher-order questions were defined as those incorporating additional intermediary steps, such as identifying a diagnosis, but subsequently requiring evaluative or analytical tasks to give the correct answer. For example, a higher-order question may present a clinical vignette and, instead of asking for the most likely diagnosis, may request the next best step of management or another clinical feature of the most likely diagnosis. This classification scheme was based on similar systems used in the setting of medical standardized examinations.^7^ Classification of questions was blinded, without prior knowledge of ChatGPT’s or GPT-4’s answers to the question.

**Table 1:** Example of First-Order vs. Higher-Order Multiple Choice Questions.

| Patient Case | Possible Questions |
| --- | --- |
| An 8-year-old girl presents with café-au-lait spots, Lisch nodules, and scoliosis. | **First-Order Question:** What is the most likely diagnosis for this case?<br>**A)** Churg-Strauss syndrome<br>**B)** Neurofibromatosis 1<br>**C)** Neurofibromatosis 2<br>**D)** Pfeiffer syndrome<br>**E)** Tuberous sclerosis |
|  | **Higher-Order Question:** What is another clinical finding associated with the most likely diagnosis in this case?<br>**A)** Subependymal giant cell astrocytoma<br>**B)** Bilateral vestibular schwannoma<br>**C)** Port-wine stain (nevus flammeus)<br>**D)** Tram-track cortical calcifications<br>**E)** Optic glioma |
| A patient who was previously GCS 15 returns from CT scan in the ER with closed eyes, no verbal output, and only withdrawing to noxious stimulation. | **First-Order Question:** Which of the following is her current Glasgow Coma Scale (GCS) score?<br>**A)** 3<br>**B)** 6<br>**C)** 8<br>**D)** 10<br>**E)** 15 |
|  | **Higher-Order Question:** Which of the following is the next most appropriate step in management?<br>**A)** IV Lorazepam<br>**B)** Place an EVD<br>**C)** Obtain an EEG<br>**D)** Obtain an MRI<br>**E)** Intubate |

Sample questions written by the authors to illustrate classification distinction between first-order and higher-order multiple choice questions.

All analyses were performed using R Version 4.1.2 (Foundation for Statistical Computing, Vienna, Austria). Linear regression was used to evaluate the associations between category-level scores. Chi-squared, Fisher’s exact, and univariable logistic regression tests were used to query differences in performance. Statistical significance was assessed at *P*<0.05. This study followed Strengthening the Reporting of Observational Studies in Epidemiology (STROBE) reporting guidelines.

## Results

### Performance of ChatGPT and GPT-4 on Neurosurgery Written Boards

On the SANS ABNS Self-Assessment Exam 1, ChatGPT (GPT-3.5) and GPT-4 achieved scores of 73.4% (367/500, 95% confidence interval [CI]: 69.3-77.2%) and 83.4% (417/500, 95% CI: 79.8-86.5%), respectively, against the user average of 73.7% (95% CI: 69.6-77.5%; **Table 2** and **Figure 2**). Question bank users and both LLMs exceeded the 2022 ABNS written board exam’s passing threshold of 69%. While scores between ChatGPT and question bank users were equivalent (*P*=0.963), GPT-4 outperformed both (both *P*<0.001).

**Table 2:** Performance of Question Bank Users, ChatGPT (GPT-3.5), and GPT-4 by Topic Area.

| Question Category | Questions | User Average | GPT-3.5 Performance | GPT-4 Performance | P-Value |  |  |
| --- | --- | --- | --- | --- | --- | --- | --- |
|  |  |  |  |  | 3.5 vs. User | 4 vs. User | 3.5 vs. 4 |
| Overall | 500 | 73.7% | 367/500 (73.4%) | 417/500 (83.4%) | 0.963 | <0.001 | <0.001 |
| Functional | 21 | 65.9% | 17/21 (81.0%) | 20/21 (95.2%) | 0.450 | 0.044 | 0.341 |
| Fundamentals | 9 | 73.1% | 8/9 (88.9%) | 9/9 (100.0%) | 1.000 | 0.471 | 1.000 |
| Neuropathology | 12 | 63.6% | 7/12 (58.3%) | 8/12 (66.7%) | 1.000 | 1.000 | 1.000 |
| Neuroradiology | 62 | 73.2% | 30/62 (48.4%) | 32/62 (51.6%) | 0.008 | 0.022 | 0.858 |
| Other General | 38 | 74.8% | 34/38 (89.5%) | 38/38 (100.0%) | 0.171 | 0.003 | 0.123 |
| Pain | 15 | 69.3% | 8/15 (53.3%) | 11/15 (73.3%) | 0.710 | 1.000 | 0.450 |
| Pediatrics | 56 | 69.8% | 41/56 (73.2%) | 45/56 (80.4%) | 0.852 | 0.285 | 0.502 |
| Peripheral Nerve | 14 | 64.3% | 14/14 (100.0%) | 14/14 (100.0%) | 0.041 | 0.041 | 1.000 |
| Spine | 92 | 79.5% | 75/92 (81.5%) | 86/92 (93.5%) | 0.878 | 0.011 | 0.026 |
| Trauma | 64 | 77.0% | 51/64 (79.7%) | 56/64 (87.5%) | 0.830 | 0.167 | 0.340 |
| Tumor | 67 | 66.0% | 50/67 (74.6%) | 62/67 (92.5%) | 0.976 | 0.021 | 0.035 |
| Vascular | 50 | 74.5% | 32/50 (64.0%) | 36/50 (72.0%) | 0.356 | 0.953 | 0.520 |

Performance of ChatGPT on 500 mock neurosurgery board examination questions, classified by subspecialty. Differences in performance were queried with chi-squared and Fisher's exact tests.

**Figure 2:**
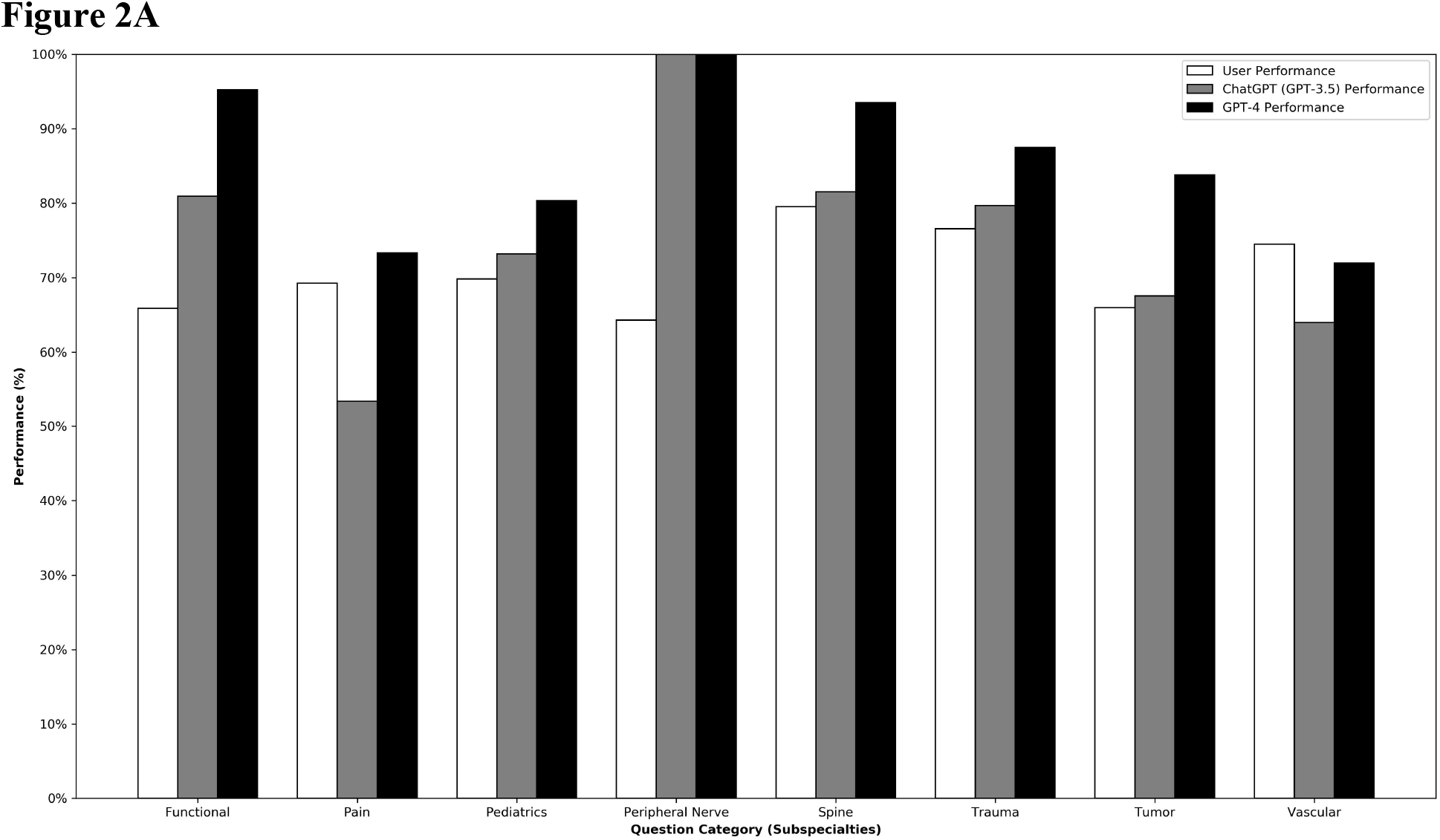
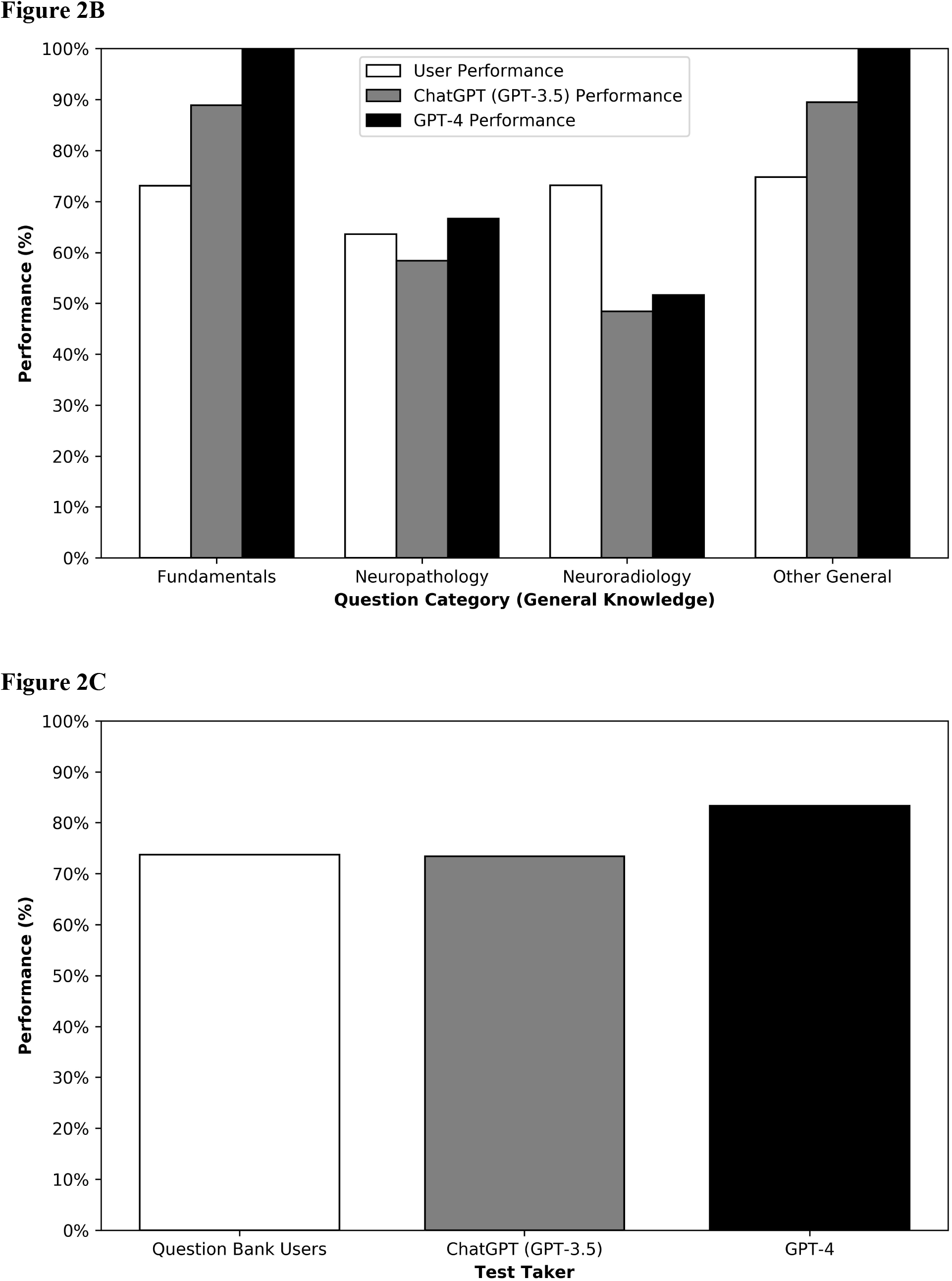
Performance of Question Bank Users, ChatGPT (GPT-3.5), and GPT-4. Histograms comparing performance of question bank users, ChatGPT, and GPT-4 by question category. **A:** Performance by neurosurgical subspecialty. **B:** Performance by categories assessing general neurosurgical knowledge. **C:** Overall performance by question bank users, ChatGPT, and GPT-4.

Of twelve question categories, GPT-4 outperformed users in every category, but performed comparably to ChatGPT in three categories (Functional, Other General, and Spine). For Tumor questions, GPT-4 outperformed both users (92.5% vs. 66.0%, *P*=0.021) and ChatGPT (92.5% vs. 74.6%, *P*=0.035). However, for Neuroradiology questions, users scored significantly higher than both ChatGPT (73.2% vs. 48.4%, *P*=0.008) and GPT-4 (73.2% vs. 51.6%, *P*=0.022). ChatGPT’s category-level performance was correlated with that of GPT-4 (r^2=0.870, *P*<0.001; **Figure 3**). However, category-level performance for question bank users was not correlated with that of ChatGPT (r^2=0.006, *P*=0.808) or GPT-4 (r^2=0.006, *P*=0.809).

**Figure 3:**
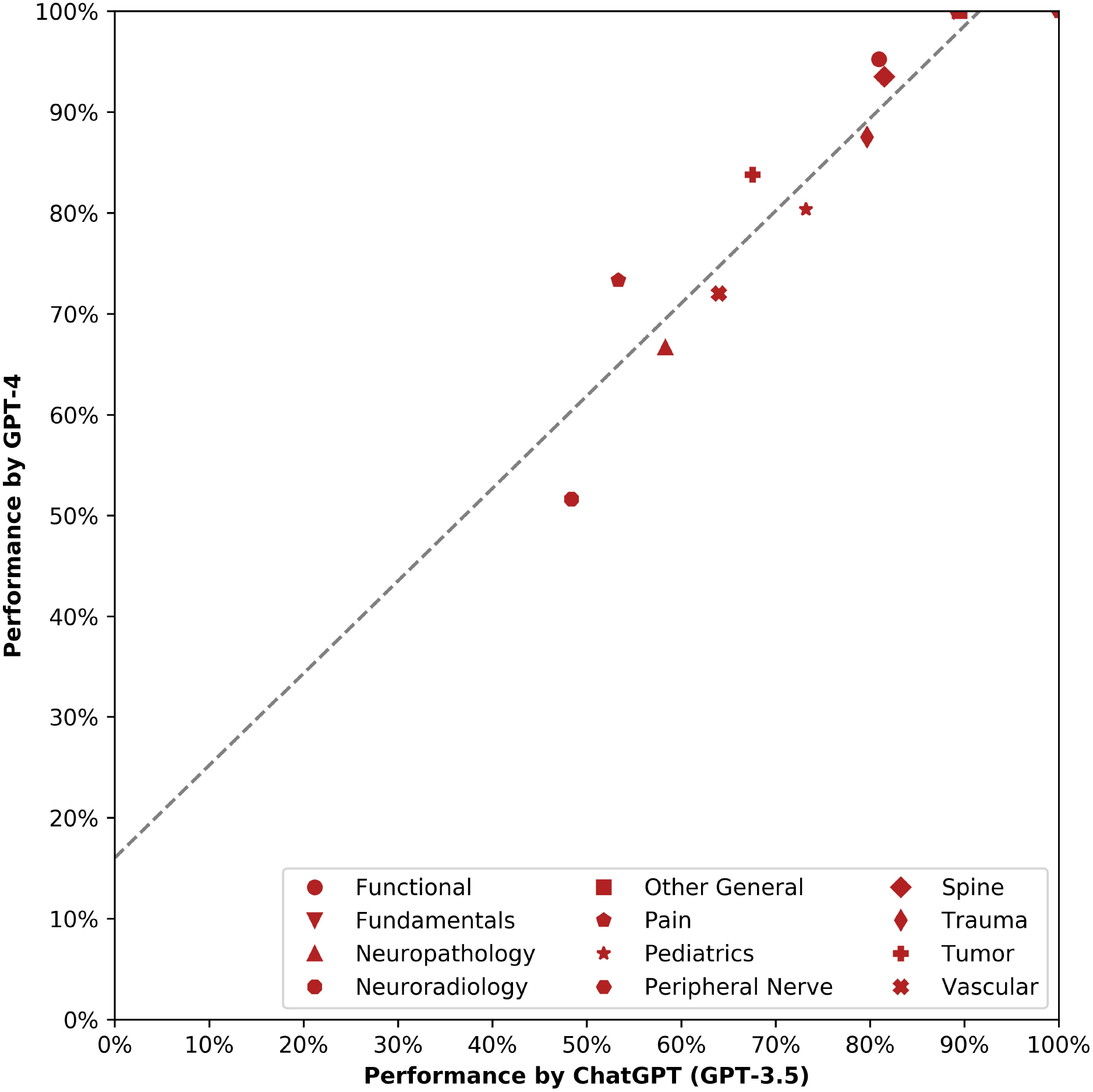
Association Between Category-Level Performance of ChatGPT (GPT-3.5) and GPT-4. Scatter plot of category-level performance for ChatGPT (x-axis) and GPT-4 (y-axis). Dotted line plots significant positive linear association between the two variables.

### Association of Question Characteristics with LLM Performance

For ChatGPT, incorrectly answered questions had a higher average word count (mean=32.2 vs. 27.9, *P*=0.025), and increased question length (odds ratio [OR]=0.89 of answering a question correctly per +10 words). 7.4% (n=37) questions were identified as requiring higher-order problem-solving. For these questions, ChatGPT was significantly less likely to return the correct answer (OR=0.40, *P*=0.009; **Figure 1E**). Nevertheless, ChatGPT did occasionally answer first-order questions incorrectly, such as failing to properly calculate a Glasgow Coma Scale (GCS) score (**Figure 1C**).

GPT-4 correctly answered all 367 questions answered correctly by ChatGPT as well as 37.6% (50/133) of the questions that ChatGPT answered incorrectly. Unlike Chat-GPT, word length (*P*=0.150) and higher-order problem-solving (*P*=0.080) were not associated with poorer accuracy by GPT-4. These characteristics were also not predictive of which questions, among those answered incorrectly by ChatGPT, were answered correctly by GPT-4. **Figure 4** depicts representative questions answered incorrectly by ChatGPT but answered correctly by GPT-4, particularly with a focus on neuroanatomy (Figure 4A-C). Additionally, GPT-4 performed an accurate GCS calculation for the question previously answered incorrectly by ChatGPT (**Figure 4D**).

**Figure 4:**
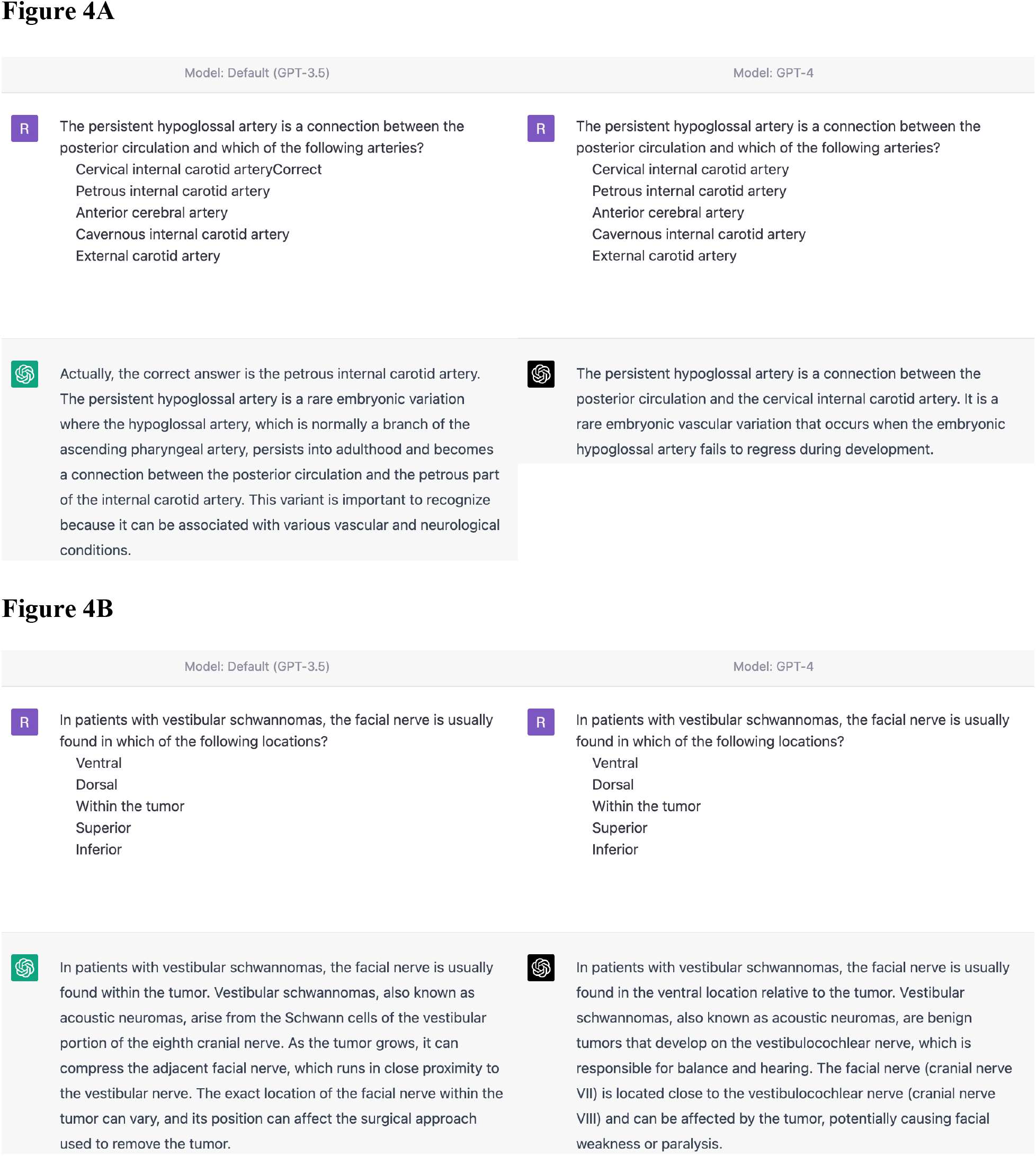
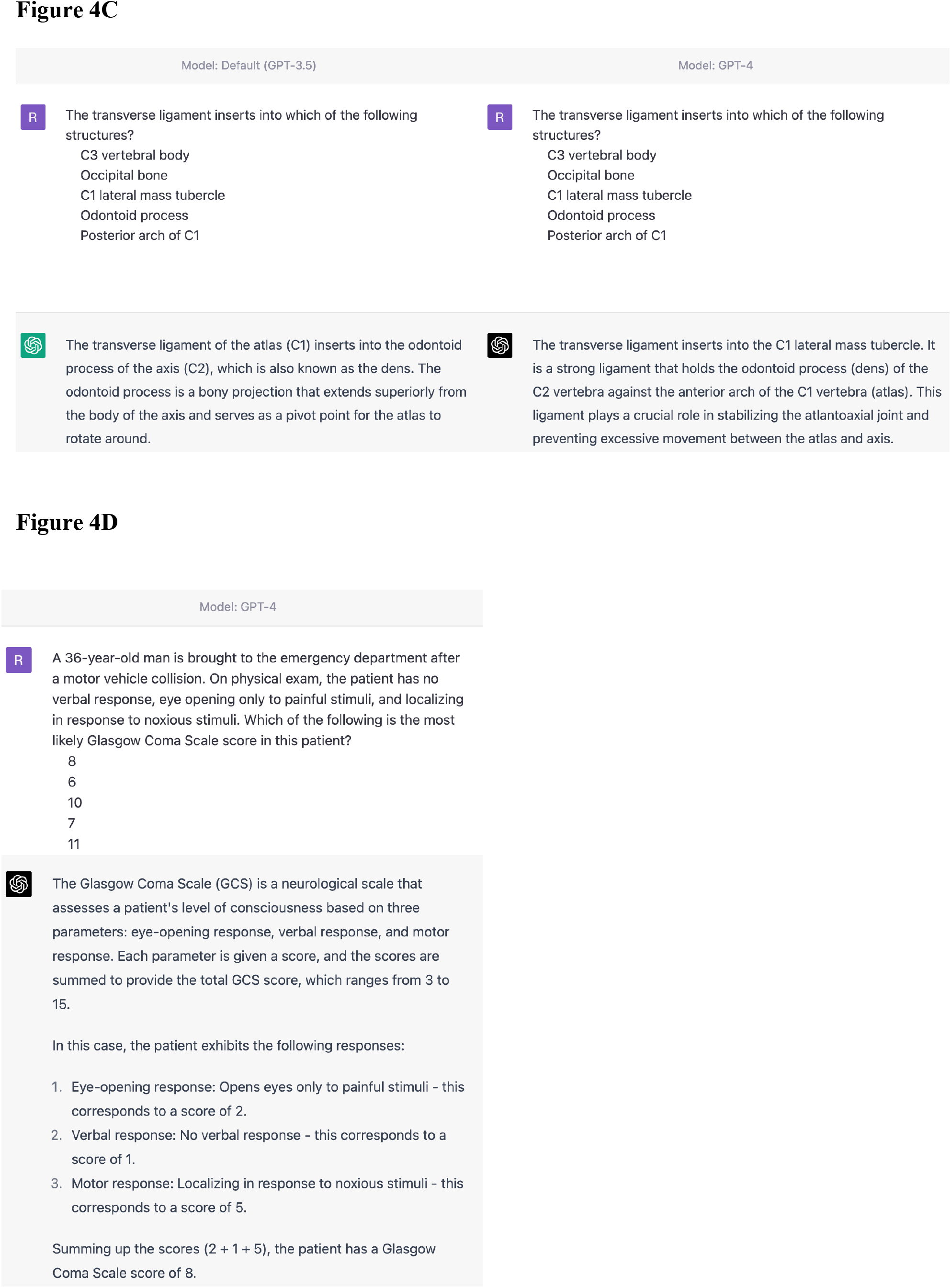
Representative Questions Answered Incorrectly by ChatGPT and Correctly by GPT-4. Screenshots of four questions answered by incorrectly by ChatGPT but answered correctly by GPT-4. All images are attributed to ChatGPT or GPT-4 (OpenAI; San Francisco, CA). **A-C:** Three questions involving neuroanatomy previously answered incorrectly by ChatGPT but correctly by GPT-4. **D:** Question involving calculation of a Glasgow Coma Scale score, which was previously answered incorrectly by ChatGPT, as demonstrated in **Figure 1C**.

### Performance on Imaging-Based Questions

There were 111 (22.2%) questions that included images (e.g., neuroradiology) that could not be entered into ChatGPT or GPT-4 at the time of this study. ChatGPT and GPT-4 declined to answer 21 and 20 image-based questions, respectively, due to lacking sufficient context (these were marked as incorrect), but attempted answers for the remaining questions. ChatGPT answered 49.5% (55/111) of image-based questions correctly, which was significantly poorer than its performance on non-imaging-related questions (80.2%, *P*<0.001). Moreover, GPT-4 answered a majority (56.8% or 63/111) of imaging-related questions correctly, which was also lower than performance on non-imaging-related questions (91.0%, *P*<0.001). Overall, scores on these image-based questions were not significantly different between ChatGPT and GPT-4.

## Discussion

The study evaluated the performance of ChatGPT (GPT-3.5) and GPT-4 on a mock neurosurgery written board examination, revealing that both AI models (with scores of 73.9% and 83.9%, respectively) exceeded the passing threshold, with GPT-4 outperforming both ChatGPT and human test-takers. Interestingly, ChatGPT’s score here was considerably higher than the model’s performance on the USMLE Step Examinations, where accuracy did not exceed 65%. Potential explanations for this finding include the more hyperspecialized nature of the content base for neurosurgical examinations or differences in question styles, such as the lengthier vignette format of USMLE questions. Notably, ChatGPT exhibited lower accuracy on longer questions and those involving higher-order problem-solving. In contrast, GPT-4 did not exhibit the same limitations, demonstrating enhanced ability to process lengthier and more syntactically complex inputs, and improved ability to navigate multiple steps of problem-solving for high-order questions.

Interestingly, GPT-4 performed best on the two topics where humans performed worst: functional neurosurgery and peripheral nerve surgery. This could reflect underexposure of human test-takers to these topics and/or something more nuanced about the types or structure of the questions themselves. Both ChatGPT and GPT-4 answered 14/14 questions correctly in the peripheral nerve section, which may be reflective of commonly tested and defined relationships inherent to this subject matter. While many anatomy-related questions can be considered first-order recall, it is important to highlight that three-dimensional anatomic relationships can often be rather complex with numerous surrounding structures. As depicted in **Figure 4A-C**, GPT-4 appeared to have improved performance on questions pertaining to neuroanatomy and spatial relationships as compared to ChatGPT. This may be due to GPT-4’s improved knowledge, reasoning, or a combination of both. In the future, it will be important to fully characterize the extent of GPT-4’s knowledge of neuroanatomy and potential shortcomings related to complex spatial relationships, which is of particular concern to surgical subspecialties.

A key constraint in both ChatGPT’s and GPT-4’s ability to answer neurosurgery board questions was our inability to incorporate imaging data. Despite this limitation, both models managed to answer a majority of imaging-related questions, albeit with lower accuracy than non-imaging-based questions. Given the progressive refinement of deep learning architectures for computer vision and medical imaging, such as Inception,^8^ it is possible that this limitation may be addressed with future AI models, such as the upcoming multimodal input functionalities of GPT-4. Future work testing LLM performance on board examination questions could also incorporate figure descriptions generated using image-to-text models.

The written boards examination was created as a benchmark and developmental requirement for neurosurgeons-in-training as a part of the broader two-part boards certification process. In a similar manner, it is important to thoroughly validate LLM’s before graduating towards widespread use, particularly within medicine. Our study serves as an initial benchmark of ChatGPT and GPT-4’s performance on an exam designed to “validate” human neurosurgical knowledge. However, this raises the interesting question of whether a test designed for humans is the best means to fully evaluate LLMs. Although correctly answered by GPT-4, a question requiring calculation of GCS score, a fairly straightforward cognitive task for any trainee, was missed by ChatGPT; this suggests there may be similar “blind-spots” yet to be discovered in GPT-4 and other LLMs. Therefore, as we integrate LLMs into clinical practice, we must strive to identify their knowledge- and reasoning-based shortcomings. Utilizing the performance of LLMs on standardized tests as a proxy to assess these capabilities may be one approach to ascertain such an understanding. The multiple-choice testing approach has the advantage of assessing performance in a straightforward, fully objective manner, but notably does not accurately mimic potential real world use of LLM’s, acting as a “copilot” for providers, where open-ended questions are inputted in various clinical situations. For example, a multiple-choice approach may not adequately assess the extent of the known phenomenon where these models might confabulate or “hallucinate” responses during more open-ended questioning.^3^

The rapid progression from ChatGPT to GPT-4, which was released only four months after its predecessor, and the clear improvements in subspecialty medical knowledge and reasoning we observed here highlight the critical need for neurosurgeons to remain knowledgeable and up-to-date about fast-evolving AI systems. As knowledge continually deepens in all areas of medicine, including neurosurgery, the value of good decision-making based on increasingly specialized and esoteric information becomes ever more critical. ChatGPT’s and GPT-4’s passing performance on neurosurgery board examinations suggests that with additional training, fine-tuning, and added multi-modal capability, AI assistance may — sooner than many might have thought even a couple of years ago — soon contribute meaningfully to medical practice, even for subspecialties like neurosurgery.

## Conclusion

Two general LLMs, ChatGPT and GPT-4, achieved passing scores on a mock 500-question neurosurgical written board examination, with GPT-4 significantly outperforming ChatGPT and question bank users. Greater question word length and incorporation of higher-order problem-solving was associated with poorer accuracy for ChatGPT, but not GPT-4. It is paramount for neurosurgeons to remain knowledgeable and up-to-date about these rapidly evolving AI systems and their potential applications to clinical medicine.

## Data Availability

Due to the proprietary nature of the dataset used for this study (Self- Assessment Neurosurgery Exams American Board of Neurological Surgery Self-Assessment Exam 1), the authors are unable to post the raw data used for the analysis. However, the authors are able to share any collected data (ex. word count, question classification, ChatGPT responses, etc.) on request to other investigators who have access to this self-assessment exam. Code used for this study's analyses is available to download from a public GitHub repository.

https://github.com/oliverytang/chatgpt_neurosurgery

